# Detection of the omicron variant in international travellers and their family contacts in India

**DOI:** 10.1101/2021.12.27.21268429

**Authors:** Varsha A. Potdar, Pragya D. Yadav, Kavita Lole, Sarah Cherian, Jayanti Shastri, Bharati Malhotra, Sumit Bharadwaj, Alpana Razdan, M L Choudhary, Nivedita Gupta, Priya Abraham, NIC team, Bioinformatics team, BSL4 team, The Indian SARS-CoV-2 Genomics Consortium (INSACOG)

## Abstract

**Highlights:** With the emergence of the Variant of Concern, omicron (B.1.1.529), India has enhanced genomic surveillance in international travellers. The omicron variant was detected in 59 cases from different States; 40 from Maharashtra, 17 from Rajasthan and one each from Gujrat and Tamil Nadu. The positive cases and their contacts were asymptomatic and genomic surveillance could identify two clusters, one from Maharashtra and another from Rajasthan.

---

The first confirmed case of the new SARS-CoV-2 Variant of Concern (VOC) was reported on November 24th, 2021 [1]. The WHO’s Technical Advisory Group on SARS-CoV-2 Virus Evolution designated the Pangolin lineage B.1.1.529; Nextstrain 21K (omicron) as VOC on November 26. [1]. When compared to the Wuhan virus, the variant spike protein comprises 30 amino acid changes, three small deletions and one short insertion, 15 in the receptor binding domain (residues 319–541). [2] Its genome also features a number of changes and deletions. The furin cleavage site contains three mutations in this variant that could further increase SARS-CoV-2 infectivity [3.].

Many of the sequences submitted by South Africa, Australia, and Canada lack the identifying mutations of B.1.1.529 (omicron). To integrate all of these variations in the B.1.1.529 lineage, two sub-lineages were identified: 21K (omicron) or BA.1 and 21L (BA.2). 21K (omicron) is of concern because to its high frequency of spike gene mutations. The spike protein of the B.1.1.529 parent lineage viz. 21M (omicron), has only 21 amino-acid mutations, with the BA.1 (omicron) sub-lineage having 12 9 distinct mutations and BA.2 having 6 mutations (T19I, V213G, S371F, T376A, D405N, and R408S) and a 9 nucleotide loss at position 21633-21641 leading to A27S/L24S mutation. The BA.2 lacks the S: H69-and S: V70-deletion that causes “S-gene drop out” or S gene target failure (SGTF) in TaqPath RT PCR assays that monitors BA.1 (omicron) [4].

As of 21 December 2021 78 countries shared 18,264 Omicron genome sequences with GISAID [5, 6]. The Indian government revised its overseas entry criteria after reports of the new SARS-CoV-2 variant (B.1.1.529; omicron). The COVID-19 test protocol requires that samples from travellers from risk nations be sent to the Indian SARS-CoV-2 Consortium on Genomics (INSACOG) network for genetic testing. The ICMR-National Institute of Virology, Pune, being a part of the network, thus sequenced clinical specimens from overseas travellers to India. Laboratories under INSACOG also shared the raw sequencing data with ICMR-NIV, Pune, which analysed and reported the sequencing results. All referenced samples during November-December 2021 were tested for SARS-CoV-2 RT-PCR. Previously published techniques (Ref 7, 8) were used to sequence the genome using the Oxford Nanopore Minion, Ion Torrent S5 and Illumina MiniSeq NGS platforms. The MagMAXTM Viral Pathogen Nucleic Acid Isolation Kit was used to isolate RNA from viral infections (Thermo Fisher Scientific, USA). Library creation and quantification followed the sequencing platform. The midnight technique was employed for Oxpford Nanopore. Commander S/w version 1.6.2 was used to sequence the complete genome. Wuhan-HU-1 was used for reference-based genome assembly. The sequences were submitted to GISAID.

A total, 1902 samples were referred for omicron testing till December 17th, 2021. 1630 came from overseas travellers, while 272 came from friends and family contacts. The states of Maharashtra, Gujarat, Tamil Nadu, and Port Blair referred the samples, with majority from Maharashtra. The referred cases had median age of 38 years (IQR 28-53 years). The referred sample comprised 823 females (43.2%). RRT-PCR detected SARS CoV-2 in 177 samples. With the procedure described above, 111 samples were processed for whole genome sequencing (WGS). Additionally, 164 samples were processed for S gene detection using the Taq Path assay and 116 samples had S gene detection, whereas 48 had SGTF. Among 116 S gene detected cases; 6 samples were detected as omicron, whereas among 48 SGTF, 22 were omicron.

We detected 29 (26.1%) as the omicron variant, 73 (65.8%) as delta and delta sub-lineages, 4 (3.6%) as other lineages and 5 (4.5%) were sequencing failures. Additionally, 30 omicron cases were identified from the sequencing analysis of raw data submitted by SMS Jaipur and Public health lab, Chennai. To summarise, 33 (55.9%) overseas travellers and 26 (44.1%) contacts reported 59 omicron incidents. The novel VOC omicron was phylogenetically analysed along with known VOCs alpha, beta, gamma, and delta, randomly selected from GISAID, preferably from India

The final dataset included 55 ICMR-NIV sequences, 7 omicron variants reported from India, and 20 VOCs. MAFFT version 7.45 was used for multiple sequence alignment. The Maximum Composite Likelihood distance approach was used to build a neighbour-joining (NJ) tree in MEGA v6 (Ref 9, 10). As seen in Fig. 1, all the sequences were of omicron BA.1 lineage except for a single sequence of lineage BA.2.

**Fig. 1.**
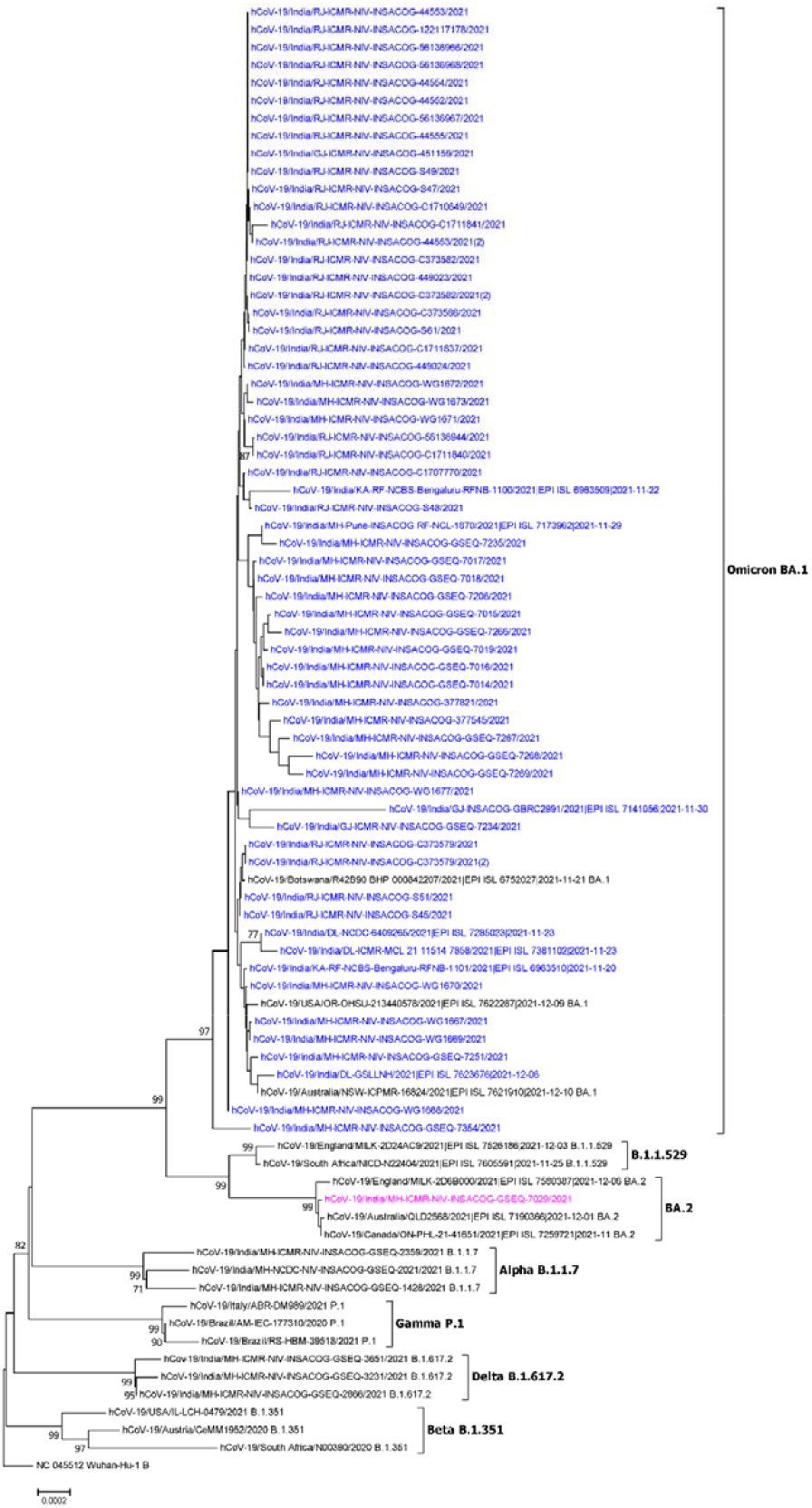
Neighbor-Joining (NJ) tree of SARS CoV 2 depicting Variants of Concern (VOC) viz. Alpha, Beta, Gamma, Delta and Omicron (blue colour)

This study discovered two distinct family clusters in which overseas returnees infected close family members (cluster-1 and cluster-2). In cluster-1, eight members of family were found. Three were index cases, while the others were contacts. The index case returned from Nigeria after testing negative for COVID-19 at Nigeria airport. Later, family members in India tested positive for omicron (Figure-2a). The whole family reported a COVID-19 infection history in August 2020.

**Figure-2a.**
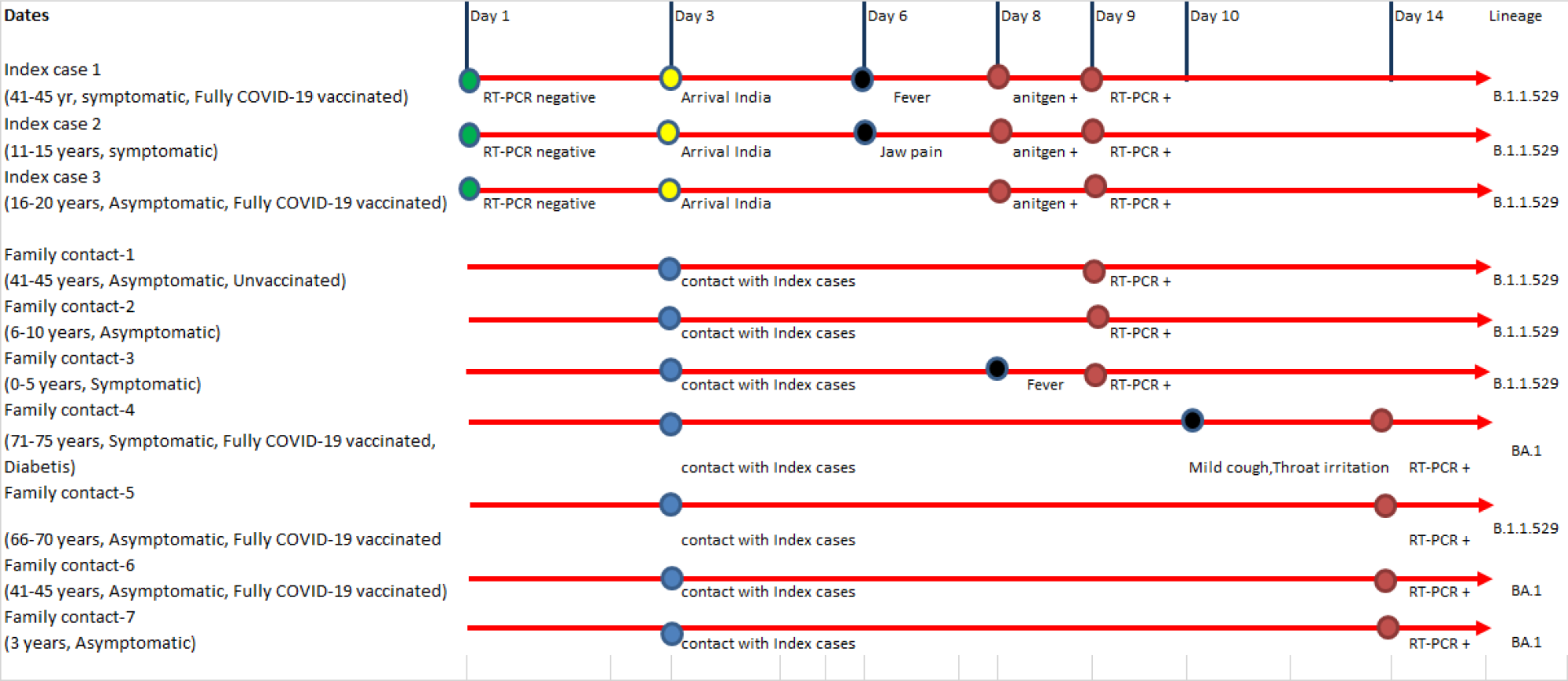
Family cluster-1

The cluster-2, in which five members of a family were infected within a four-hour period of brief exposure by the index cases. Index cases were tested RT-PCR negative at Johannesburg airport (Figure-2b) In September 2020, all these index cases and family members had COVID-19.

**Figure-2b.**
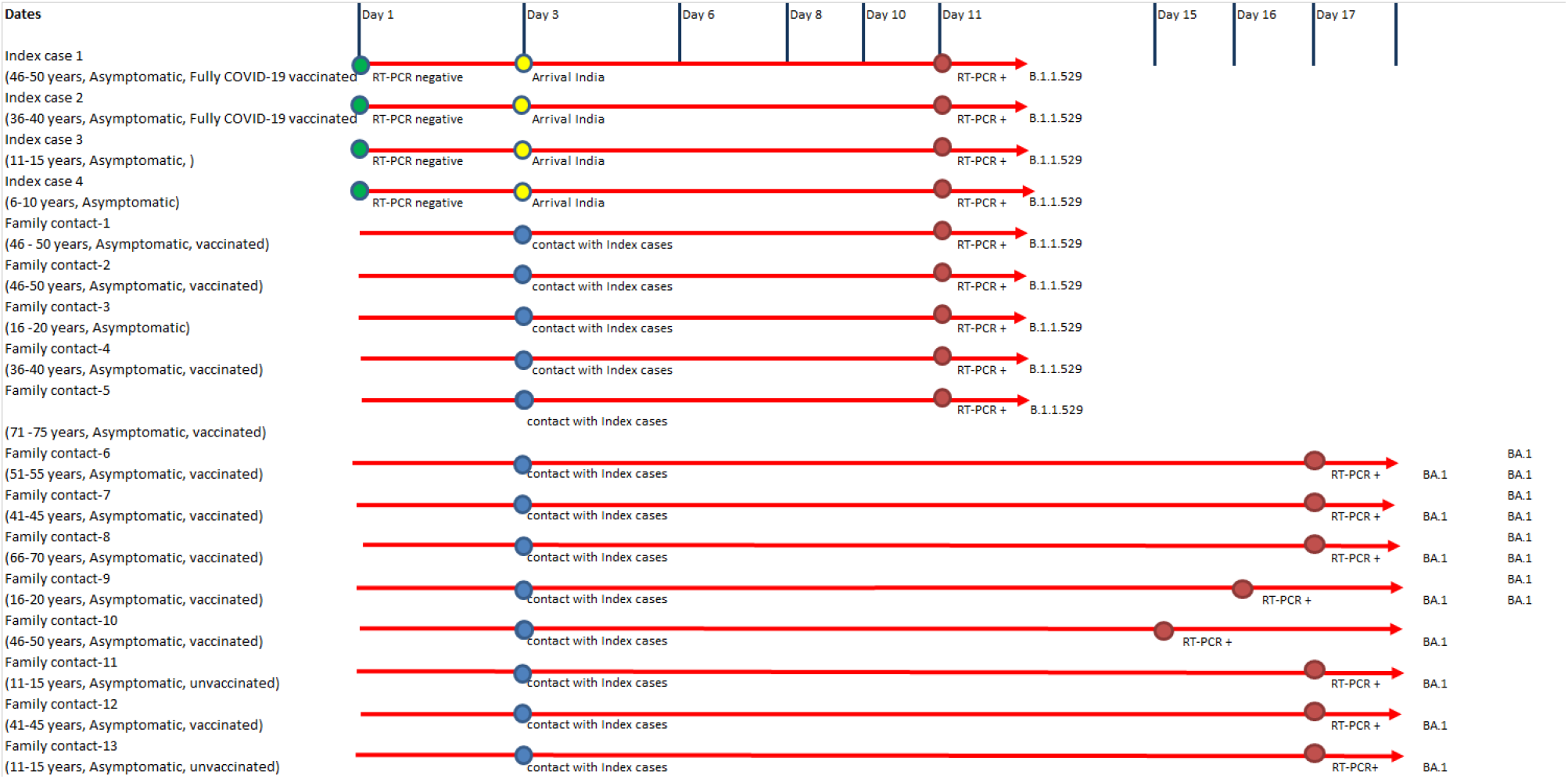
Family cluster-2

Omicron was readily spread through close contact, with a three-to-six-day incubation period. Seventeen of the omicron cases in the family cluster were vaccinated against COVID-19, while one adult was unvaccinated. None of the afflicted family members required medical assistance or hospitalisation. It was confirmed that all the contact cases were SARS CoV-2 re-infections.

Several laboratories have reported the use of S gene detection as a screening tool for identification of probable omicron variants. However, the SGTF is also reported in other VOCs like alpha; hence cautioning the use of S gene target. Our study showed 50% of cases of SGTF had the omicron variant and that this test can thus be used as a flag for this variant where sequencing confirmation is pending. In this article, two omicron-infected family clusters are presented. WGS confirmed omicron infection in members of this family cluster. It was discovered incidentally under the screening programme for foreign returnees and their close contacts. Fortunately, the three symptomatic family members recovered completely. The INSACOG hub at Pune further sequenced 1665 community samples during November from Maharashtra and no omicron variant was detected. As of now, omicron positivity is reported only among international travellers and their contacts. Family members of persons who have been diagnosed with COVID-19 should be closely monitored and tested for virus infection, even if they exhibit no symptoms.

## Ethical approval

The study was approved by the Institutional Biosafety Committee and Institutional Human Ethics Committee of ICMR-NIV, Pune, India under the project titled ‘Molecular epidemiological analysis of SARS-COV-2 circulating in different regions of India’.

## Data Availability

All data produced in the present work are contained in the manuscript

## Author contributions

VAP, PDY, SC, KL, JS, BM, AR accessed and verified the data. VAP, PDY, SC, SB contributed to the analysis and manuscript preparation. VAP was the study coordinator, PA, MLC, KL, SC reviewed the manuscript. VAP was responsible for overall supervision of the study and review of the final paper.

## Acknowledgement

Dr Pradip Awate and State IDSP team. Mumbai Municipal Corporation, Pune Munciple Corporation and Pimpri Chinchwad Municipal Corporation for sample referral.

## List of INSACOG Consortium Members

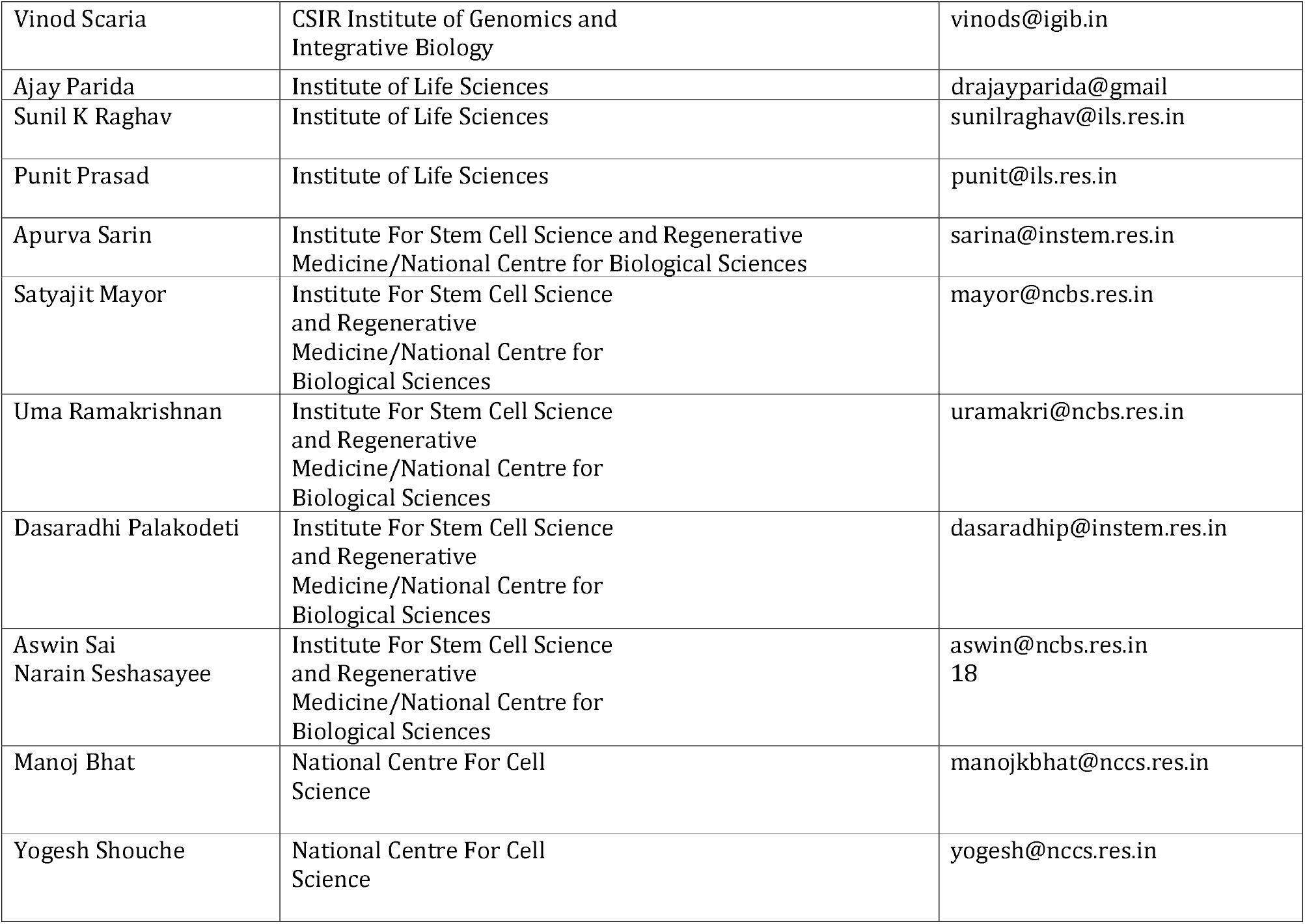
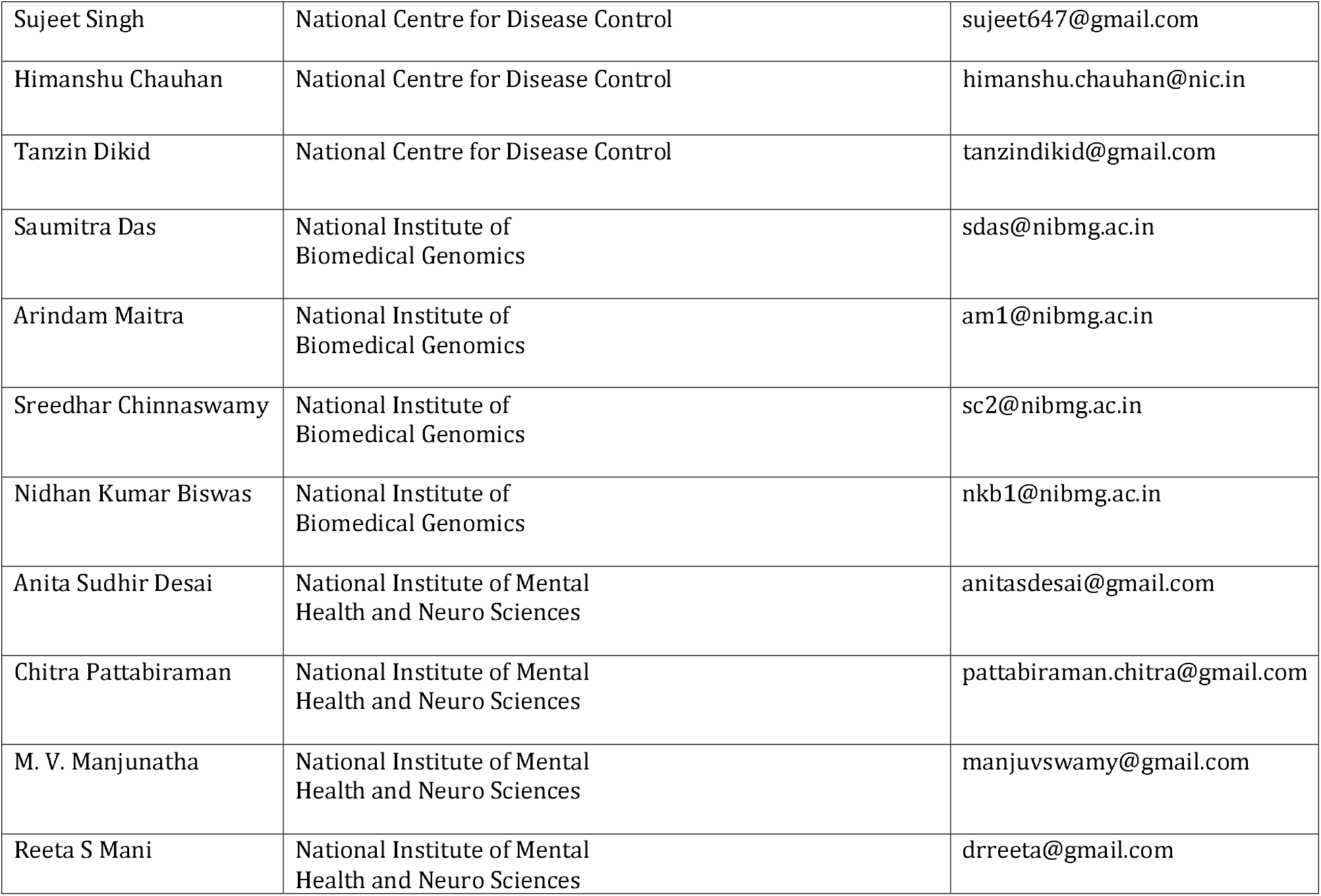
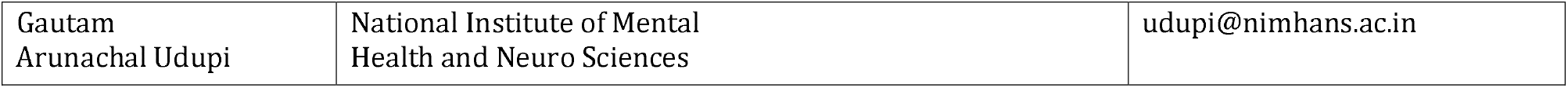

